# A Genome-Informed Functional Modeling Approach to Evaluate the Responses of Breast Cancer Patients to CDK4/6 Inhibitors-Based Therapies and Simulate Real-World Clinical Trials

**DOI:** 10.1101/2023.05.15.23289976

**Authors:** Mei Yang, Yuhan Liu, Chunming Zhang, Yi-Ching Hsueh, Qiangzu Zhang, Yanhui Fan, Juntao Xu, Min Huang, Xu Li, Jianfei Yang, Guangming Tan, Gang Niu

**Author notes:** Correspondence and requests for materials should be addressed to Dr. Guangming Tan via and Dr. Gang Niu via. These authors equally contribute to this work (Mei Yang, Yuhan Liu).

## Abstract

**PURPOSE:** Varied therapeutic responses were observed among cancer patients receiving the same treatment regimen, highlighting the challenge of identifying patients most likely to benefit from a given therapy. Here, we present an artificial intelligence-based approach, called CDK4/6 inhibitor Response Model (CRM), to address the complexity of predicting patient responses to treatment by a certain clinical scene on CDK4/6 inhibitors (CDK4/6i).

**PATIENTS AND METHODS:** To train the CRM, we transformed the genomic data of 980 breast cancer patients from the TCGA database into activity profiles of signaling pathways (APSP) by utilizing the modified Damage Assessment of Genomic Mutations (DAGM) algorithm. A scoring model was then established by random forest algorithm to classify the HR+/HER2− and HR−/HER2− breast cancer molecular subtypes by the differential APSP features between the two, which reasonably reflected the potential role played by CDK4/6 molecules in HR+/HER2− breast cancer cells. The effectiveness of CRM was then tested in a separate local patient cohort (n = 343) in Guangdong, China. Twin in-silico clinical trials (ICT) of previously disclosed clinical trials (NCT02246621, NCT02079636, NCT03155997, NCT02513394, NCT02675231) were performed to demonstrate the potential of CRM in generating concerted results as the real-world clinical outcomes.

**RESULTS:** The CRM displayed high precision in classifying HR+/HER2− and HR−/HER2− breast cancer patients in both TCGA (AUC=0.9956) and local patient cohorts (AUC=0.9795). Significantly, the scores were distinct (p = 0.025) between CDK4/6i-treated patients with different responses. Breast cancer patients from different subtypes were grouped into five distinct populations based on the scores assigned by the CRM. From twin ICT, the CRM scores reflected the differential responses of patient groups to CDK4/6i-based therapies.

**CONCLUSION:** The CRM score showed not only a robust association to clinically observed CDK4/6i responses but also heterogenetic responses across subtypes. More than half of HR+/HER2+ patients may be benefited from CDK4/6i-based treatment. The CRM empowered us to conduct ICT on different types of cancer patients responding to CDK4/6i-based therapies. These findings showed the potential of CRM as the companioned ICT to guide CDK4/6i application in the clinical end. CRM-guided ICT could be a universal method to demonstrate drug sensitivity to various patients.

## Introduction

In clinical practice, patients often respond differently to the same drug, making it challenging to identify the most suitable population for a particular treatment. Traditional methods for identifying biomarkers to predict drug sensitivity and resistance have been found more and more ineffective, and the underlying mechanisms behind patient responses remain poorly understood^1^. As artificial intelligence (AI) methods continue to advance in biomedical research, they offer great potential for exploring the mechanisms of drug response^2, 3^. Here, we present a study focused on CDK4/6 inhibitor (CDK4/6i)-based therapies, demonstrating the ability of AI methods to identify patients’ sensitivity to this treatment.

CDK4/6, cyclin dependent kinase 4 and 6, plays a key role in regulating the cell cycle of tumor cells^4–6^. Therefore, drugs targeting CDK4/6 have achieved success in some clinical applications, benefiting certain patient groups from CDK4/6 inhibitor (CDK4/6i)-based therapy^7–9^. For instance, CDK4/6 inhibitors have been approved by FDA for treating adult patients with hormone receptor (HR)-positive, human epidermal growth factor receptor 2 (HER2)-negative advanced or metastatic breast cancer in combination with endocrine therapy as initial treatment in postmenopausal women or in men^10^. Despite the successes, it is evident that the application conditions of CDK4/6i-based therapies are relatively narrow, limited to specific conditions inside breast cancer and lung cancer. Currently, researchers are exploring further on CDK4/6 inhibitors in basic research and clinical aspects^1, 11–17^.

In the past, many explorations of biomarkers have been conducted to detect CDK4/6i-sensitive or resistant patients^18–20^. However, the sensitivity evaluation method for CDK4/6i-based therapies is still an unmet clinical need, and no clear clinical practice standard can predict whether patients will respond to the treatment based on any single feature, such as CCND1, CCNE1 and p16 loss^21–27^. It was suggested that multiple drug resistance mechanisms are involved in the CDK4/6i resistance, and the population covered by the single gene biomarkers is far less than the proportion of patients with actual drug resistance^28, 29^.

The other current sensitivity screening methods, such as scattered single nucleotide polymorphisms (SNPs) of pharmacogenomics, and simple association analysis like genome-wide association studies (GWAS), mainly rely on surface-level characteristics and also fall short in revealing the drug sensitivity^30–32^. The fundamental reason for the inefficiency in patient screening lies in the lack of a clear understanding of how tumors respond to CDK4/6i-based therapies, leading to inconsistent patient stratification based on response outcomes.

Through clinical observations, it has been found that some therapies, such as endocrine therapy and CDK4/6 inhibitors, are effective for patients with HR+/HER2− breast cancer but not for those with HR−/HER2− breast cancer^33, 34^. Therefore, it is promising to establish drug-sensitivity screening methods by comparing the systematic differences of HR+/HER2− and HR−/HER2− breast cancer in molecular biological characteristics to understand the underlying mechanism of drug sensitivity. Notably, CDK4/6 inhibitors exhibit different mechanisms of action on the two breast cancer subtypes, leading to significant differences in clinical efficacy. In HR+/HER2− breast cancer, CDK4/6i-based therapies directly inhibits tumor cell growth, resulting in significant improvements in patient outcomes^35^. However, the efficacy of CDK4/6i-based therapies for patients with triple-negative breast cancer (HR−/HER2−) is relatively weak, with only a small number of patients benefiting from CDK4/6 inhibitors as a pre-treatment to protectively inhibit bone marrow-derived cells, ultimately leading to the continuation of immune system functions after other treatments^36, 37^. Hence, it can be inferred that CDK4/6 inhibitors directly inhibit the growth of tumor cells, which is the common mechanism for a higher proportion of HR+/HER2− subtypes to benefit from CDK4/6i-based therapies.

In our previous study on the etiology of germline genomes in breast cancer patients, we proposed a potential solution for analyzing functional genomics differences^38^. By mapping germline rare coding variants (gRCVs) onto a quantitative set of signaling pathway profiles using the Damage Assessment of Genomic Mutations (DAGM) approach, we can easily model the functional patterns of cells driven by germline genomes in these patients. This method can not only distinguish between HER2-negative and positive patients but can also construct a scoring model to accurately predict the relative risk of HER2-negative breast cancer in female individuals, even those with “apathogenic” gene, like wild-type BRCA1/2. Building on this approach, we can modify the algorithm to analyze how somatic mutations drive deterministic changes in cell function, providing a methodological basis for distinguishing between different pathological tumor types.

We developed a CDK4/6 inhibitor response model (CRM) based on the genomic profiles of breast cancer patients from The Cancer Genome Atlas (TCGA) and an independent local patient cohort in Guangdong, China. By converting the spectrum of somatic rare mutations to activity profiles of signaling pathways (APSP) using DAGM, the systematic differences between HR+/HER2− and HR−/HER2− subtypes in functional biology level can be analyzed and used for training the CRM. The CRM can distinguish HR+/HER2− and HR−/HER2− subtypes, which reflects the sensitivity to CDK4/6 inhibitor therapies.

Due to limited clinical efficacy data and the interests in the simulation capability of in-silico clinical trial (ICT), we employed ICT to validate the CRM, which also interprets the effectiveness of ICT to simulate real-world clinical trials. We established digital patient profiles based on TCGA as the twin of clinically enrolled patients to simulate responses to CDK4/6 inhibitors. The results suggest the CRM was able to reflect the differences in treatment efficacy across trials, which can probably identify patients suitable for the targeted therapies. We propose the CRM as a potential precision medicine tool for better patient stratification and the CRM-based ICT method as a powerful approach with clinical utility.

## Methods

### Patient Cohort and Data Collection

In this study, we analyzed the Whole Exome Sequencing (WES) data of 980 breast cancer patients from The Cancer Genome Atlas (TCGA) and 541 breast cancer patients from Guangdong, China. The two cohort was categorized into four subtypes based on the expression of ER (estrogen receptor), PR (progesterone receptor), and HER2, including HR+/HER2− type (Luminal A and Luminal B1 type, n=454 in TCGA, n=206 in local cohort), HR+/HER2+ type (Luminal B2 type, n=58 in TCGA, n=92 in local cohort), HR-/HER2+ type (HER2-enriched type, n=33 in TCGA, n=82 in local cohort), and HR−/HER2− type (TN type, n=127 in TCGA, n=137 in local cohort). The classification of breast cancer was based on Goldhirsch’s research on 2013^39^. WES data was obtained from peripheral blood and tumor lesions of the patients to facilitate a comprehensive analysis of somatic mutations in breast cancer genomes.

### CRM establishment, verification and optimization

To establish a prediction model for CDK4/6 inhibitor-based therapy, the baseline characteristics of four patient groups in TCGA were compared. The adapted DAGM algorithm was used to analyze the clonal somatic activity profiles of signaling pathways (sAPSP) based on the genomic data of patients’ tumor tissues and peripheral blood samples. The sAPSP were represented as a list of quantitative measurements of signaling pathways, indicating their activation and inhibition status. The APSP characters were defined as differences in the mean of the APSPs between subtypes, with the Z score used to assess significant differences. APSP characters with an absolute Z score ≥3 were considered as differential features between HR+/HER2− and HR−/HER2− subtypes.

Based on the differential features selected above as model parameters, CRM was established using the random forest method, which generates a score between 0 and 1. The data of HR+/HER2− and HR−/HER2− breast cancer patients in TCGA were used as the training set. Patients with a score closer to 1 have tumor profiles closer to that of HR+/HER2− breast cancer patients and are suitable for CDK4/6i-based therapies. Conversely, patients with a score closer to 0 have tumor profiles closer to that of HR−/HER2− breast cancer patients and are likely not sensitive to CDK4/6i-based therapies. The classification ability of CRM was self-validated by leave-one-out cross validation (LOOCV)^40^.

To further validate the CRM, the local WES data of HR+/HER2− and HR−/HER2− subtypes were used as testing sets. The CRM’s performance was further improved by combining TCGA and local patient cohort as training data, leading to an enhanced model in distinguishing patients. The optimized model was verified by LOOCV again.

### Validation of the CRM score and CDK4/6i-efficacy

To validate the linkage between the CRM scores and drug efficacy, 13 female patients with pathologically diagnosed breast cancer were recruited from the Guangdong local database. The patients, aged between 26 and 72 years old, were tested for molecular typing by immunohistochemistry. The 13 patients were all diagnosed with HR+/HER2− type breast cancer. All patients received CDK4/6i (Palbociclib)-based combination therapy, which included one or more drugs in fulvestrant, anastrozole, zoledronic acid, letrozole, exemestane, everolimus, and Norad. These patients were at different stages of treatment, including first-line, second-line, or multi-line post-treatment, and the shortest PFS (progression-free survival) among them was 2 months.

As defined by objective response rate (ORR) in RECIST^41^, patients with complete response (CR) and partial response (PR) were considered as responders of CDK4/6i, whereas patients with stable disease (SD) and progressive disease (PD) were considered as non-responders of CDK4/6i. The CRM was applied to evaluate these responders and non-responders to check if the CRM could accurately reflect the response differences.

### The CRM prediction on other breast cancer subtypes

To further explore the predictive ability of the CRM, breast cancer patients from TCGA and local databases were scored based on their cancer subtype classification. Specifically, HR+/HER2− patients were further divided into Luminal A type (Ki-67 < 15%) and Luminal B1 type (Ki-67 >= 15%) based on the Ki-67 index measured in local clinical detection and chosen from the study of PAM50 and Claudin-low (CLOW) molecular subtypes for TCGA patients^42^. The CRM scores were calculated for each subtype, especially HR+/HER2+ and HR-/HER2+, and their distribution characteristics were analyzed to predict the response to CDK4/6i-based therapies in different breast cancer subtypes.

### In-silico clinical trials

#### Clinical Trials Selection

Five clinical trials were chosen for this study, which employed CDK4/6 inhibitors-based therapy to treat breast cancer and non-small cell lung cancer (NCT02246621 was assigned as No.1, NCT02079636 as No.2, NCT03155997 as No.3, NCT02513394 as No.4, NCT02675231 as No.5). The No.1 to No.4 trials were selected by distinct outcomes observed clinically, and the No.5 trial was chosen to demonstrate the possible correlation between the CRM scores and drug efficacy. The basic information of the five selected clinical trials is presented in Table 1.

**Table 1:** Basic information of selected clinical trials^44, 45, 47, 49, 59^

| Serial number | NCT number | Drug name | Trial results | Conditions | Drug in test | Drug control |
| --- | --- | --- | --- | --- | --- | --- |
| 1 | NCT02246621 | Abemaciclib | Succeeded | Advanced HR+HER2-<br>breast cancer | Abemaciclib+NSAI<br>(Anastrozole/Letrozole,<br>endocrine therapy) | Placebo + NSAI<br>(Anastrozole/Letrozole,<br>endocrine therapy) |
| 2 | NCT02079636 | Abemaciclib | Not<br>succeeded | Stage IV NSCLC | Abemaciclib+Multiple<br>single agent options<br>(pemetrexed,<br>gemcitabine, or<br>ramucirumab) | none |
| 3 | NCT03155997 | Abemaciclib | Succeeded | Early high-risk, node-<br>positive, HR+/HER2-<br>breast cancer | Abemaciclib +<br>Standard adjuvant<br>endocrine therapy | Standard adjuvant<br>endocrine therapy |
| 4 | NCT02513394 | Palbociclib | Not<br>succeeded | Early HR+/HER2-<br>breast cancer | Palbociclib + Standard<br>adjuvant endocrine<br>therapy | Standard adjuvant<br>endocrine therapy |
| 5 | NCT02675231 | Abemaciclib | Succeeded | HR+/HER2+ locally<br>advanced or metastatic<br>breast cancer | Abemaciclib +<br>Trastuzumab +<br>Fulvestrant | Trastuzumab +<br>standard<br>chemotherapy |

#### Simulated patients screening

To simulate patients enrolled in clinical trials, 950 and 1051 patients with complete clinical information from the TCGA breast cancer and lung cancer database (TCGA-BRCA, TCGA-LUAD, TCGA-LUSC) were included. The specific clinical information of eligible patients in TCGA was selected to screen out digital twins that closely resemble clinical trial patients as shown in Supplementary Table 1.

#### Results comparison

The CRM scores of simulated patients in each trial were calculated and plotted to illustrate their distributions. These simulated results were compared across trials and with real-world clinical observations. Additionally, for the No.5 clinical trial, the scores of simulated patients were preliminarily screened to observe if they can accurately reflect the clinical outcomes of patients.

## Results

### General concept of establishing the CRM to evaluate patients’ sensitivity to CDK4/6i-based therapies

As shown in Figure 1, in most cases, the efficacy of CDK4/6 inhibitors (CDK4/6i) for breast cancer patients is tightly correlated with molecular subtypes, which are classified by the status of hormone receptors (HR) and human epidermal growth factor receptor 2 (HER2). More than one pathological condition in HR+/HER2− type of patients are approved to use CDK4/6i-based therapies as standard treatment, while patients with HR−/HER2− breast cancer have less chances to benefit from CDK4/6i-based therapies. Therefore, the differential features of cellular function between HR+/HER2− and HR−/HER2− breast cancers were able to reflect the working mechanism of CDK4/6i. To establish CRM, we used an AI-based approach to firstly transform the genomic information of the two types of breast cancer to activity profiles of signaling pathways (APSP), and then selected differential APSP between the two to train a scoring model by machine learning. The CRM was capable for identifying the differential responses of breast cancer patients to CDK4/6i-based therapies by the CRM-given score.

**FIG 1:**
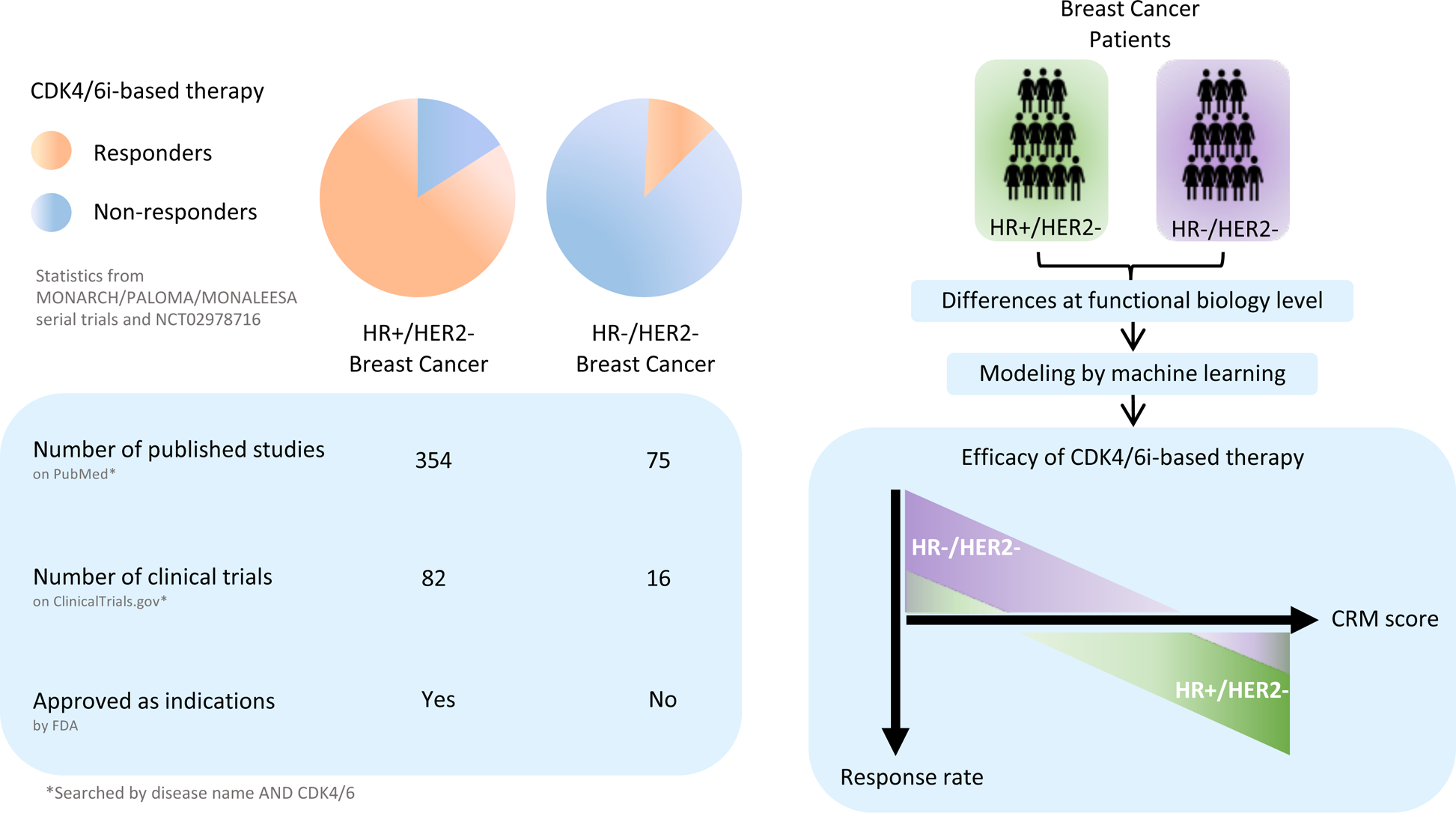
The general concept of establishing the CRM to evaluate patients’ sensitivity to CDK4/6i-based therapies. Varied responses to CDK4/6i-based treatment were found between HR+/HER2− and HR−/HER2− breast cancers, as high response rate observed in HR+/HER2− but not in HR−/HER2−^25, 33, 44, 52–58^. The number of related research also displayed diversity because of the differential drug efficiency. Up to Mar 1, 2023, the number of CDK4/6i studies in HR+/HER2− breast cancer was greatly larger than that of HR−/HER2− breast cancer. CDK4/6i approved indications were also gathered in HR+/HER2− breast cancer. Based on these findings, the CDK4/6i response model (CRM) was established by machine learning method on differential features between HR+/HER2− and HR−/HER2− breast cancers in the functional biology level. This model was able to reflect the distinct responses to CDK4/6i in HR+/HER2− and HR−/HER2− breast cancer patients.

### Development of CRM in HR+/HER2− and HR−/HER2− breast cancer patients

#### Characterization of TCGA-BRCA patients

The baseline characteristics of the four types of breast cancer patients in TCGA were well-balanced across the subtypes (Table 2), with the majority of patients aged from 41 to 70 (72% in HR+/HER2− and HR+/HER2+, 93% in HR-/HER2+, 73% in HR−/HER2−) and most of them being female. The patient population in all subtypes included three or more races, such as Asian, black or African American, and white. Most of the patients were diagnosed at stage II according to staging information.

**Table 2:** The baseline characteristics of the CRM training set and other TCGA-BRAC patients

|  | HR+/HER2-<br>n=454 |  | HR+/HER2+<br>n=58 |  | HR-/HER2+<br>n=33 |  | HR-/HER2-<br>n=127 |  |
| --- | --- | --- | --- | --- | --- | --- | --- | --- |
|  | n | % | n | % | n | % | n | % |
| **Gender** |  |  |  |  |  |  |  |  |
| Female | 451 | 99.34% | 56 | 96.55% | 33 | 100.00% | 127 | 100.00% |
| Male | 3 | 0.66% | 2 | 3.45% | 0 | 0.00% | 0 | 0.00% |
| **Age** |  |  |  |  |  |  |  |  |
| 20-30 | 4 | 0.88% | 1 | 1.72% | 0 | 0.00% | 0 | 0.00% |
| 31-40 | 31 | 6.83% | 8 | 13.79% | 1 | 3.03% | 13 | 10.24% |
| 41-50 | 88 | 19.38% | 9 | 15.52% | 10 | 30.30% | 35 | 27.56% |
| 51-60 | 112 | 24.67% | 18 | 31.03% | 10 | 30.30% | 41 | 32.28% |
| 61-70 | 128 | 28.19% | 14 | 24.14% | 7 | 21.21% | 24 | 18.90% |
| 71-80 | 66 | 14.54% | 5 | 8.62% | 4 | 12.12% | 10 | 7.87% |
| 81-90 | 25 | 5.51% | 3 | 5.17% | 1 | 3.03% | 4 | 3.15% |
| **Ethnicity** |  |  |  |  |  |  |  |  |
| American Indian or Alaska Native | 0 | 0.00% | 0 | 0.00% | 1 | 3.03% | 0 | 0.00% |
| Asian | 22 | 4.85% | 4 | 6.90% | 10 | 30.30% | 7 | 5.51% |
| Black or African American | 53 | 11.67% | 7 | 12.07% | 3 | 9.09% | 38 | 29.92% |
| White | 332 | 73.13% | 37 | 63.79% | 18 | 54.55% | 76 | 59.84% |
| Not reported | 47 | 10.35% | 10 | 17.24% | 1 | 3.03% | 6 | 4.72% |
| **AJCC pathologic stage** |  |  |  |  |  |  |  |  |
| I | 92 | 20.26% | 7 | 12.07% | 2 | 6.06% | 22 | 17.32% |
| II | 248 | 54.63% | 37 | 63.79% | 21 | 63.64% | 83 | 65.35% |
| III | 101 | 22.25% | 14 | 24.14% | 8 | 24.24% | 18 | 14.17% |
| IV | 13 | 2.86% | 0 | 0.00% | 1 | 3.03% | 1 | 0.79% |
| X | 6 | 1.32% | 0 | 0.00% | 0 | 0.00% | 0 | 0.00% |
| Not reported | 2 | 0.44% | 0 | 0.00% | 1 | 3.03% | 3 | 2.36% |
| **Pathologic type at primary diagnosis** |  |  |  |  |  |  |  |  |
| Infiltrating duct carcinoma, NOS | 307 | 67.62% | 51 | 87.93% | 32 | 96.97% | 111 | 87.40% |
| Lobular carcinoma, NOS | 102 | 22.47% | 4 | 6.90% | 0 | 0.00% | 6 | 4.72% |
| Infiltrating duct and lobular carcinoma | 13 | 2.86% | 0 | 0.00% | 0 | 0.00% | 0 | 0.00% |
| Infiltrating duct mixed with other types of carcinoma | 10 | 2.20% | 1 | 1.72% | 0 | 0.00% | 1 | 0.79% |
| Others | 22 | 4.85% | 2 | 3.45% | 1 | 3.03% | 9 | 7.09% |

#### CRM for Classifying HR+/HER2− and HR−/HER2−

The differential APSP features between HR+/HER2− and HR−/HER2− subtypes in TCGA were used to establish the CRM for predicting response to CDK4/6i-based therapies by random forest method (Figure 2a). As shown in Figure 2b, cell cycle G2M checkpoint regulation was excessively activated in HR−/HER2− subtype, while substantial inhibition appeared in HR+/HER2− subtype. In contrast, iCOS-iCOSL signaling in T helper cells was largely suppressed in HR−/HER2− breast cancer but stimulated in HR+/HER2− breast cancer.

**FIG 2:**
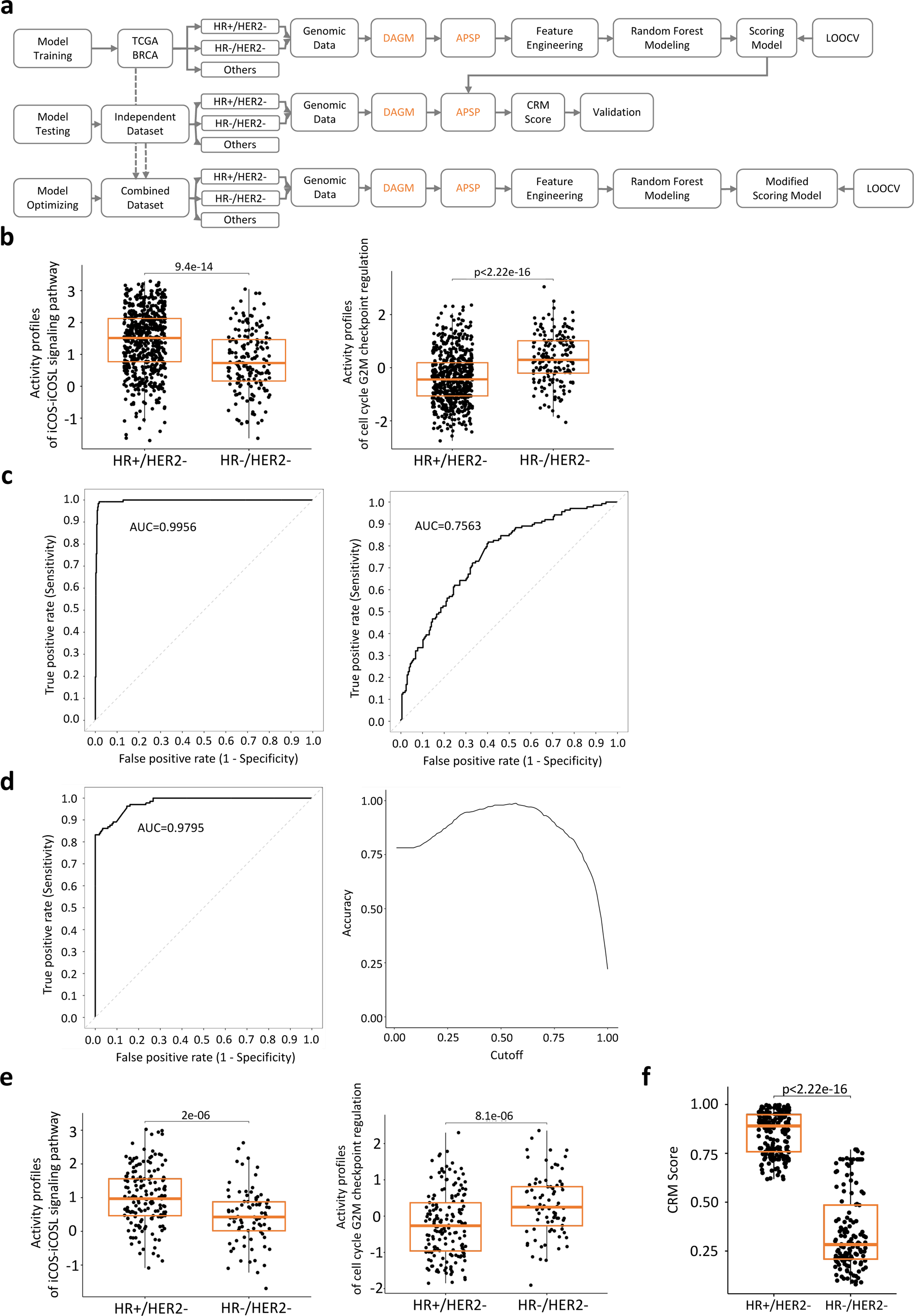
The CRM for classification of HR+/HER2− and HR−/HER2− subtypes based on genomic data. (A) Schematic of the model training, testing, and optimization workflow. (B) DAGM analysis reveals significant differences in cell cycle G2M checkpoint regulation and iCOS-iCOSL signaling pathway between HR+/HER2− and HR−/HER2− subtypes in TCGA. (C) ROC-AUC curves of the TCGA data-trained CRM for classifying TCGA (left) and local (right) HR+/HER2− and HR−/HER2− patients. (D) ROC-AUC curves of the optimized CRM for classifying local HR+/HER2− and HR−/HER2− patients by leave-one-out cross validation (LOOCV). The accuracy curve demonstrates the model’s excellent separation ability. (E) DAGM analysis of local patient dataset reveals significant differences in cell cycle G2M checkpoint regulation and iCOS-iCOSL signaling pathway between HR+/HER2− and HR−/HER2− subtypes. (F) The CRM scores show a clear distinction between HR+/HER2− and HR−/HER2− subtypes in the local patient dataset.

The CRM’s effectiveness was initially validated in the training set by the leave-one-out cross-validation (LOOCV) method, and the AUC for distinguishing HR+/HER2− from HR−/HER2− subtypes was 0.9956 (Figure 2c).

#### CRM generalization ability confirmed in an independent patient cohort

The analysis on the independent patient cohort in Guangdong, China shows that the CRM can effectively distinguish between HR+/HER2− and HR−/HER2− subtypes of breast cancer (AUC=0.7563, accuracy=70% when cutoff set to 0.6, FDR=30%). The finding confirms the generalization ability of the model in a patient cohort of different ancestries and suggest its potential usefulness in clinical settings.

#### Optimization of CRM improves classification efficiency

After combining the TCGA and Guangdong local patient data for model training, the CRM’s effectiveness in distinguishing HR+/HER2− and HR−/HER2− patients was significantly improved during the LOOCV verification process. The AUC for classifying local patients increased from 0.7563 to 0.9795 and FDR reduced to 5% after optimization (Figure 2d). Similar activated and suppressed patterns of iCOS signaling and G2M checkpoint were observed in the local patient cohort (Figure 2e). The distinction between CRM scores of HR+/HER2− and HR−HER2− patients was significant (p value < 2.22e-16, Figure 2f). The improvement indicates that the optimized CRM has a better ability to differentiate the two breast cancer subtypes from different ancestries. It suggests that the optimized CRM is more reliable and accurate in predicting the subtype of breast cancer patients, and thus can facilitate the personalized response evaluation.

### The CRM scores effectively predict response to CDK4/6i-based therapies in a retrospective analysis

In Guangdong patient database, 6 patients achieved PR, 6 were SD, and 1 was PD after receiving CDK4/6i-based therapies. A total of 22 samples, including breast and lymphoid lesions, were analyzed using the CRM for these 13 patients. The median CRM score of responding patients was 0.9181 (Figure 3a), significantly higher than that of non-responding patients (median: 0.8047, p=0.0209; MRI images from some CDK4/6i-treated patients shown in Figure 3b). The CRM scores therefore have a strong correlation with clinical outcomes when using CDK4/6i-based therapies. It demonstrates the ability of the CRM scores in evaluating the response of patients to CDK4/6i-based therapies and potential to be used as a prospective tool in the future.

**FIG 3:**
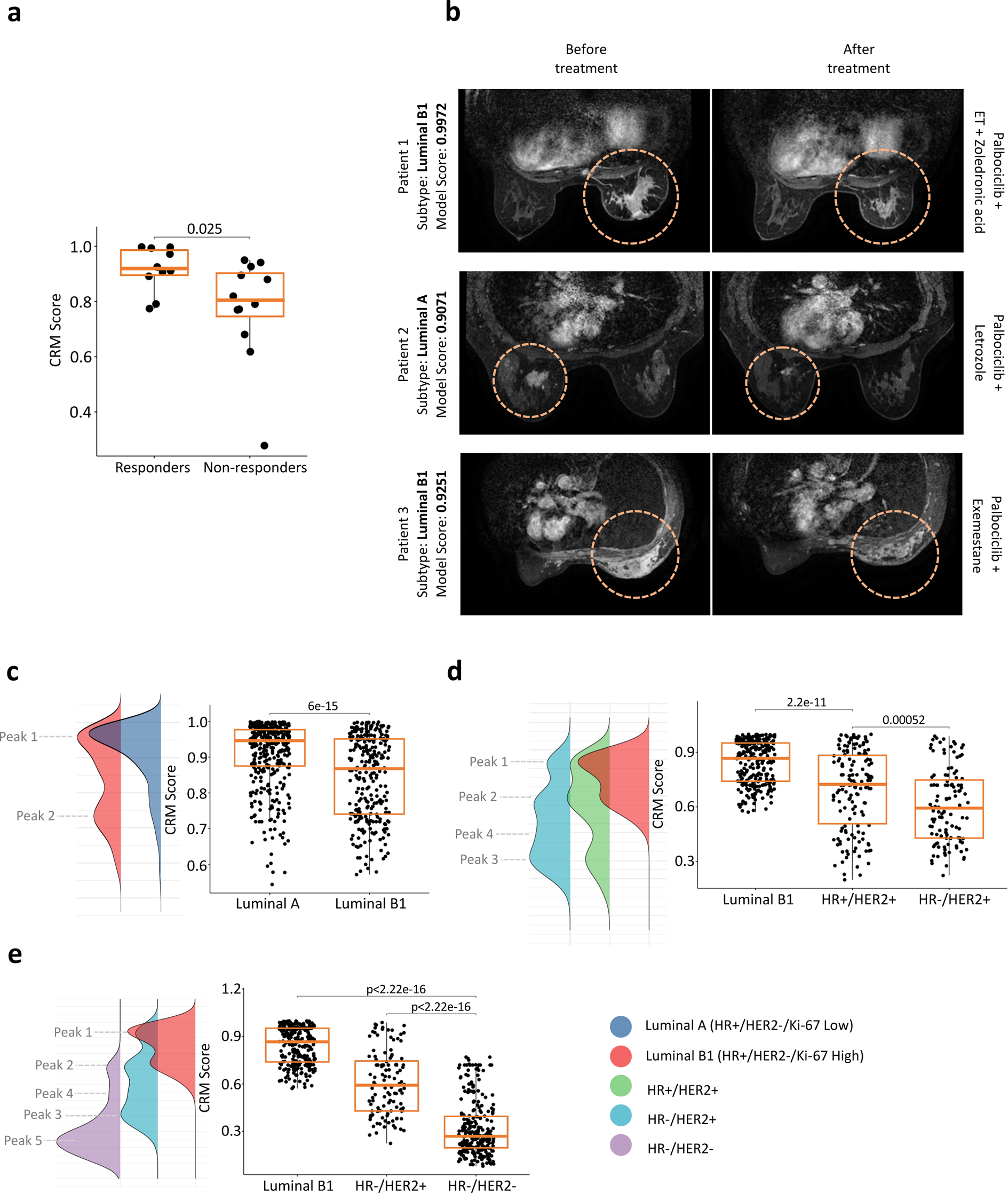
The CRM scores may reflect patient responses. (A) Boxplots show the CRM scores of responders and non-responders to CDK4/6i-based therapies, indicating significantly higher scores in the responders’ group (p value=0.017). (B) MRI data before and after using CDK4/6i-based therapies illustrate a strong correlation between CRM scores and drug efficacy. (C) Boxplots of the CRM scores for Luminal A and Luminal B1 subtypes of HR+/HER2− patients show significant variation. (D) The HER2 positive subtypes exhibit a broader pattern than the Luminal B1 subtype, with partial overlap. (E) Boxplot of HR-/HER2+ subtype scores indicate a lower score range compared with HR+/HER2+ subtype.

### Subtype-specific CRM score distribution suggests differential response to CDK4/6i-based therapies

The CRM scores of all collected breast cancer subtypes were analyzed to predict their response to CDK4/6i-based therapies. It is noted that the score distributions of the five types of patients (Luminal A, Luminal B1, HR+/HER2+, HR-/HER2+, HR-/HR2-) vary significantly and rank from high to low. Heterogeneity is observed in different types of breast cancer, illustrated as various peaks inside one distribution curve of a certain breast cancer subtype.

From CRM scores analysis, firstly, stratification of HR+/HER2− patients may be necessary when using CDK4/6i-based therapies. By combining the Ki-67 index, the highest scored HR+/HER2− patients were divided into Luminal A (Ki-67 Low) and Luminal B1 (Ki-67 High) types. Surprisingly, the CRM was able to identify the difference between Luminal A and Luminal B1 patients without incorporating the Ki-67 index in model training, suggesting that Luminal A and Luminal B1 types indeed have distinct functional biology. The scores of Luminal A type were significantly higher than those of Luminal B1 type (Figure 3c). Both Luminal A and Luminal B1 subtypes displayed two peaks (peak 1: mean=0.95, peak 2: mean=0.73), with Luminal B1 subtype having more samples distributed in peak 2 than Luminal A subtype. These results suggest that Luminal A and Luminal B1 types should be evaluated separately when assessing the sensitivity of patients to CDK4/6i-based therapies.

By observation from the score distribution of HR+/HER2+ and HR-/HER2+ subtypes, CDK4/6i-based therapies may also benefit some of the patients. The score distributions of HER2 positive patients show three distinct peaks (Figure 3d, peak 1, 2, and 3: mean=0.40). The distributions of Luminal B1 and HER2 positive HR+/HER2+ patients have a clear overlapping region. However, HR+/HER2+ subtype primarily distributes in peaks 1 and 2, while HR-/HER2+ subtype allocates more in peaks 2 and 3 (Figure 3d). The HR-/HER2+ subtype also exhibits a mild peak 4 (mean=0.55), which is also present in the HR−/HER2− subtype (Figure 3e). These findings suggest that compared to HR+/HER2+ subtypes, only a small proportion of HR-/HER2+ patients may benefit from CDK4/6i-based therapies.

Furthermore, the score comparison between HR-/HER2+ and HR−/HER2− subtypes indicates that a large proportion of HR-/HER2+ patients may be resistant to CDK4/6i-based therapies, as demonstrated by the large intersecting area of scores with HR−/HER2− subtype (Figure 3e). Three peaks are observed in the HR−/HER2− subtype distribution, which are peak 2, peak 4, and peak 5 (mean=0.24). Peak 5 is a unique pattern of HR−/HER2− subtype, also indicating the uniqueness of HR−/HER2− subtype in responding to CDK4/6i-based therapies. These features observed in the CRM score distribution could be a valuable predictor of response to CDK4/6i-based therapies.

### CRM for interpreting clinical outcomes and harnessing the potential by in-silico clinical trials

#### Insights from in-silico clinical trials on advanced-stage patients

The No.1 (NCT02246621) clinical trial was simulated following the described steps (Figure 4a) to represent a patient group with high response rate to CDK4/6i (clinically observed ORR: 55.4%, PFS: 28.2 months) ^43, 44^. The trial examined the combination of CDK4/6i (Abemaciclib) and a nonsteroidal aromatase inhibitor in postmenopausal women with no prior systemic therapy in the advanced setting. In contrast, the efficacy of CDK4/6i in stage IV non-small cell lung cancer (NSCLC) is limited (clinically observed median PFS ranging from 1.58 to 5.55 months)^45^. We then simulated the No.2 (NCT02079636) trial, which compared different combinations of CDK4/6i with other drugs, like gemcitabine, ramucirumab and pemetrexed, to treat patients with stage IV NSCLC.

**FIG 4:**
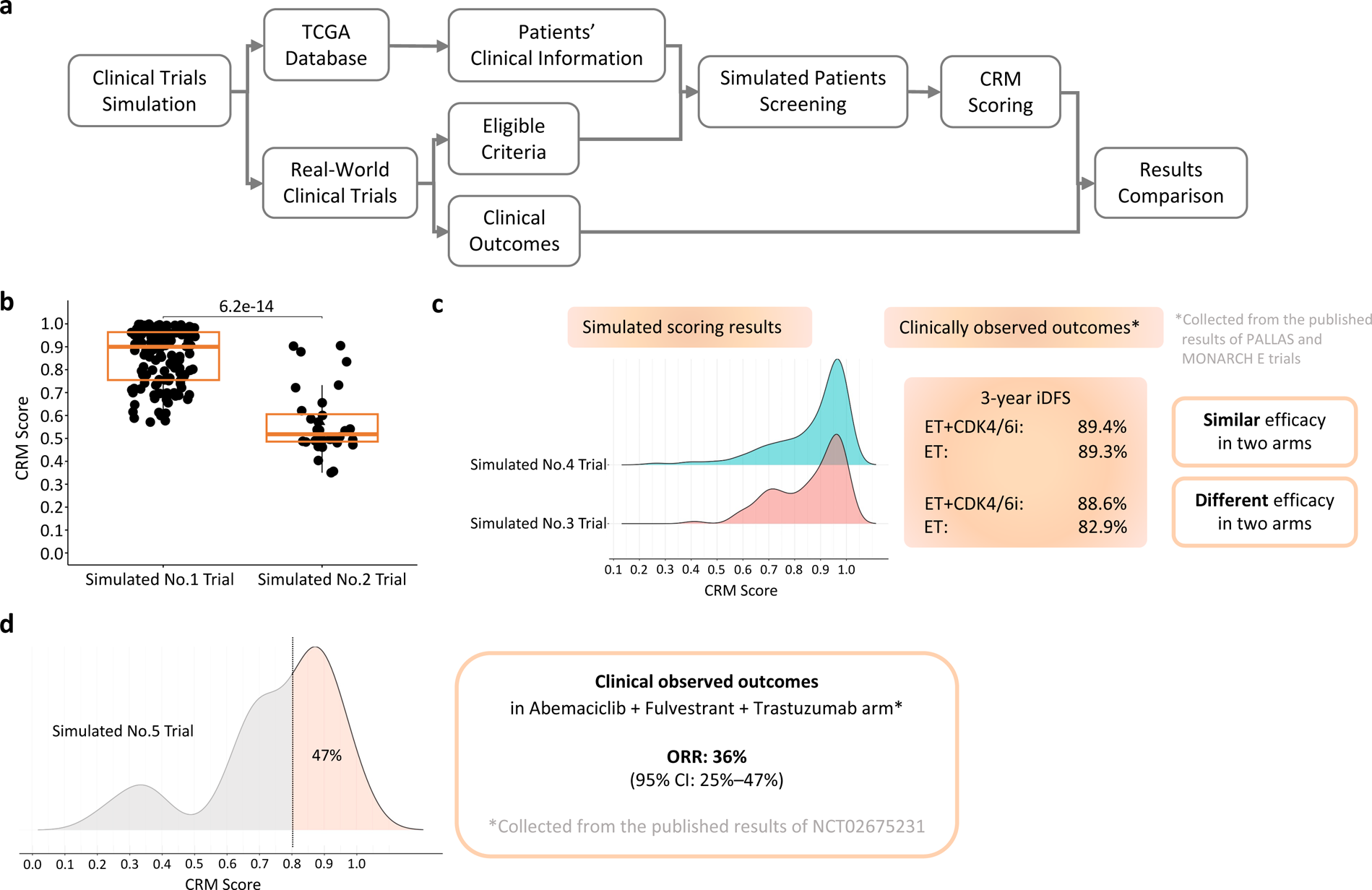
Simulation of Clinical Trials. (A) The workflow of the simulation process for clinical trials. (B) Boxplots displaying significant differences in CRM scores between simulated No.1 and No.2 clinical trials. (C) Score distributions of simulated No.3 and No.4 clinical trials exhibit notable divergence in the peak 2 region. (D) Two distinct peaks were observed in the score distribution of the simulated No.5 clinical trial. A score threshold of 0.80 is set to differentiate the two peaks, resulting in 47% of patients being identified as potential candidates for CDK4/6i-based therapies.

As depicted in Figure 4b, the CRM scores of the No.1 trial evidence a significant discrepancy compared to the No.2 trial, reflecting the response differences in these two patient groups (p value < 6.2e-14). The simulation result indicates that the CRM scores and the efficacy of CDK4/6i-based therapies are tightly correlated when evaluating advanced-stage patients.

#### In-silico simulation reveals varying efficacy of CDK4/6i-based therapies in Luminal A and Luminal B1 subtypes

The patients enrolled in the No.3 (NCT03155997) and No.4 (NCT02513394) trials are simulated with well-balanced baseline characteristics between the two groups (Supplementary Table 2). In line with our previous hypothesis, published clinical results confirmed the association between CDK4/6 inhibitors and different outcomes in patients with Luminal A and Luminal B1 subtypes. Clinical trials were conducted to evaluate whether the addition of CDK4/6 inhibitors to standard adjuvant endocrine therapy would improve efficacy in early HR+/HER2− breast cancer patients, but the outcomes of the trials were divergent. The additional benefit brought by CDK4/6i was observed in the No.3 trial but not in the No.4 trial, although the No.4 trial achieved higher iDFS rates in both arms^46, 47^. Intriguingly, the Ki-67 index may play a crucial role in the varying outcomes of the two trials^48^. Based on the No.3 trial, CDK4/6i was approved for treating Luminal B1 patients. The additional efficacy conferred by CDK4/6i is more pronounced in Luminal B1 subtype compared to Luminal A subtype, despite the better prognosis of Luminal A-type patients following treatment.

As mentioned previously, Luminal A and Luminal B1 subtypes have similar two-peak distributions, but Luminal B1 samples tend to have a greater allocation at the minor peak compared to Luminal A samples. By ICT, it is found that the CRM score distribution differs between the No.3 and No.4 clinical trials (Figure 4c, major peak: mean=0.95, minor peak: mean=0.70). The simulated No.4 trial has more samples distributed in the higher score range (major peak), while distribution of the No.3 trial leans towards the lower score range (minor peak). These results are suggested to be correlated with the better but indifferent efficacy in both arms of treatments in the No.4 trial, and the opposite observation shown in the No.3 trial. In general, when using CRM to predict the additional efficacy brought by CDK4/6i as adjuvant therapy for early breast cancer, it is easier to demonstrate efficacy differences by recruiting patients with the CRM scores in the minor peak range.

#### Simulation of trial in HR+/HER2+ breast cancer and the potential of the CRM as a companion diagnostic

The clinically observed results of the No.5 trial (NCT02675231) showed the combination of Abemaciclib, fulvestrant and trastuzumab significantly improved PFS and ORR of advanced HR+/HER2+ breast cancer patients when compared with standard chemotherapy + trastuzumab (ORR: 36% vs 16%)^49^. In the simulation, the CRM scores of the enrolled patients (n=28) were distributed as a three-peak pattern surrounding 0.87, 0.70, and 0.33, respectively (Figure 4d), indicating that the enrolled patients were heterogeneous in response to CDK4/6i-based therapies. The median score of simulated patients was 0.8181, and patients with scores larger than 0.8 (the dividing point between the higher peaks) accounted for 47% of simulated patients, falling within the 95% confidence interval of the ORR observed in the CDK4/6i, fulvestrant and trastuzumab group in the clinical study. These results suggest that the CRM has the potential for establishing companion diagnostics for CDK4/6i-based therapies by screening patients in a proper score range based on their disease and medication status.

## Discussions

In this study, we developed a novel AI approach using the adapted Damage Assessment Framework of Genomic Mutations (DAGM) algorithm to derive pathway-level quantitative information, named as activity profiles of signaling pathways (APSP), from tumor genomes. The information was used for identifying differential features between HR+/HER2− and HR−/HER2− type of breast cancers and predict response to CDK4/6i-based therapies. We trained CRM on TCGA data and successfully validated it on 343 patients from Guangdong, China. The CRM scores are strongly linked to CDK4/6i responses verified by CDK4/6i-treated patients and five patient clusters were identified by the model with differential responses. These findings also hypothesize that a large proportion of patients with HR+/HER2+ breast cancer might be benefited from CDK4/6i-based therapies. Furthermore, real-world clinical trials simulation showed CRM’s ability to manifest differences in patient responses observed in clinical practice.

The transformed functional information, which named APSP, of patients can be considered as a form of digital twins of the real patients. The ICT applied personal APSP and the CRM as a digital drug can be considered as an accurate simulation of “patients on medication” and can be easily adapted and applied in real clinical practice. As shown in ICT, simulated patients with stage IV NSCLC presented low CRM scores, corresponding with the clinical outcomes. The CRM was applied to patients with other cancer and guided the CDK4/6i-treatment in investigator-initiated research, and the current results are proven to be positive (unpublished). These findings encouraged the CRM as a potential companion diagnostic to be applied in pan-cancer treatment.

From the CRM scoring, patients with some diseases might be mostly concentrated at a high range like Luminal A breast cancer. When considering medication scheme, patients with high-score disease exhibit higher possibility for responding to CDK4/6i-based therapy and may not be required to take companion diagnostic test by CRM. For disease with heterogenetic CRM score distributions like HR+/HER2+ patients, the CRM as companion diagnostic is necessary and proper medication scheme can be assigned to suitable patients, which approach the goal of precision medication.

The CRM can be applied to screen suitable patients in different clinical stages or lines of treatment with modifications. For instance, patients with relapsed HR+/HER2− advanced breast cancer after multiple lines of therapies may possess tumors with severe malignancy, and their response rates to various treatments may be significantly lower than patients in other conditions. Thus, adjustments in the CRM score distribution pattern and screening methods may be necessary. In the MONARCH 1 (NCT02102490) clinical trial, for evaluating CDK4/6i efficacy in the heavily treated HR+/HER2− patients, the ORR for CDK4/6i was 19.7%^50^, which is lower than the outcomes of clinical trials for patients in other conditions. This result supported the hypothesis that the CRM score distribution of these patients may be concentrated at the lower range, and the threshold for screening patients in this condition might also be lower than the other conditions.

Massive attempts were carried out to expand the usage of CDK4/6i-based therapies^51^, and the CRM might provide a new prospective to this field. The CRM-like methods carry the ability to investigate the relationship between CDK4/6i-responding mechanism and the cellular mechanism facilitating tumor growth or immune response. By collecting responses to other therapies, like PARP inhibitors and anti-angiogenic therapies, we could establish models like the CRM and find if these therapies having complementary or opposing effects in anti-tumor mechanisms. Hence, the drugs could be rationally assigned to patients and boost the curative capacity of the drugs.

## Data Availability

All data produced in the present study are available upon reasonable request to the authors

## Acknowledgements

We thank all the patients who participated in this study and the medical staff who collected clinical data and samples. We also thank Guangdong Provincial People’s Hospital that provided data and clinical guidance for this study. We acknowledge the invaluable contributions of the Philrivers computational biology team that provided critical analysis and interpretation of the data. We thank West Institute of Computing Technology for offering high performance computing platform. We express our gratitude to all the colleagues who contributed to this study through their expertise, insights, and support. This work was supported by National Natural Science Foundation of China (grant no. 82072939 to M. Yang), Natural Science Foundation of Guangdong Province (grant no. 2022A1515010425 to M. Yang), Guangzhou Science and Technology Program (grant no. 202206010110 to M. Yang), Hefei National Laboratory for Physical Sciences at the Microscale (grant no. KF2020009 to G. Niu), National Natural Science Foundation of China (grant no. T2125013 to G. Tan).

## Author contributions

G. Niu and Y. Liu conceived the study. G. Niu, M. Yang, Y. Liu and Q. Zhang designed the study. M. Yang collected clinical data from patients and provided clinical guidance. G. Niu, Y. Liu, Q. Zhang, Y. Fan, J. Xu and X. Li performed data analysis. C. Zhang and G. Tan guided the application of high-performance computing. Y. Liu, M. Yang, Y. Hsueh, J. Yang and G. Niu wrote the manuscript. All authors reviewed and approved the final version of the manuscript.

