## Supplementary Table 1 for "A Genome-Informed Functional Modeling Approach to Evaluate the Responses of Breast Cancer Patients to CDK4/6 Inhibitors-Based Therapies and Simulate Real-World Clinical Trials"

### The criteria to match TCGA patients to clinical trials simulation

**a**

| NCT02246621<br>(the No.1 trial) | | NCT02079636<br>(the No.2 trial) | |
|---|---|---|---|
| **Eligible criteria of the clinical trial** | **Corresponding clinical information of TCGA patients** | **Eligible criteria of the clinical trial** | **Corresponding clinical information of TCGA patients** |
| 18 Years and older (Adult, Older Adult) | (TCGA-BRCA patients are all 18 years old and above) | 18 Years and older (Adult, Older Adult) | (TCGA-LUSC/LUAD patients are all 18 years old and above) |
| Female | gender: Female | NSCLC | Lung Squamous Carcinoma<br>OR<br>Lung Adenocarcinoma |
| HR+ | er_status_by_ihc: Positive<br>OR<br>pr_status_by_ihc: Positive | Stage IV | pathologic_stage: IV |
| HER2- | her2_fish_status: Negative<br>copy number: Less than 9<br>OR<br>her2_fish_status: [Not Evaluated]/Equivocal/Indeterminate<br>her2_status_by_ihc: Negative<br>copy number: Less than 9 | | |
| Postmenopausal | menopause_status: Post | | |
| Locally recurrent or metastatic disease | Metastatic:<br>ajcc_pathologic_m: M1<br><br>High risk of recurrence*:<br>ajcc_pathologic_n: N2/N2a/N3/N3a/N3b/N3c<br>OR<br>ajcc_pathologic_t: T3/T3a/T4/T4b/T4d<br>OR<br>PAM50 and Claudin-low (CLOW) Molecular Subtype*:<br>LumB | | |

**b**

| NCT03155997<br>(the No.3 trial) | | NCT02513394<br>(the No.4 trial) | |
|---|---|---|---|
| **Eligible criteria of the clinical trial** | **Corresponding clinical information of TCGA patients** | **Eligible criteria of the clinical trial** | **Corresponding clinical information of TCGA patients** |
| 18 Years and older (Adult, Older Adult) | (TCGA-BRCA patients are all 18 years old and above) | 18 Years and older (Adult, Older Adult) | (TCGA-BRCA patients are all 18 years old and above) |
| HR+ | er_status_by_ihc: Positive<br>OR<br>pr_status_by_ihc: Positive | ER+ and/or PR+ | er_status_by_ihc: Positive<br>OR<br>pr_status_by_ihc: Positive |
| HER2- | her2_fish_status: Negative<br>copy number: Less than 9<br>OR<br>her2_fish_status: [Not Evaluated]/Equivocal/Indeterminate<br>her2_status_by_ihc: Negative<br>copy number: Less than 9 | HER2- | her2_fish_status: Negative<br>copy number: Less than 9<br>OR<br>her2_fish_status: [Not Evaluated]/Equivocal/Indeterminate<br>her2_status_by_ihc: Negative<br>copy number: Less than 9 |
| Invasive breast cancer | histological_type: Infiltrating Ductal Carcinoma/Infiltrating Lobular Carcinoma/Infiltrating Carcinoma NOS<br>OR<br>histological_type: Other Specify/[Not Available]<br>Histology Annotations*:Invasive ductal carcinoma/Invasive lobular carcinoma/Invasive micropapillary carcinoma/Invasive carcinoma with medullary features/ Invasive carcinoma, type cannot be determined | Invasive breast cancer | histological_type: Infiltrating Ductal Carcinoma/Infiltrating Lobular Carcinoma/Infiltrating Carcinoma NOS<br>OR<br>histological_type: Other Specify/[Not Available]<br>Histology Annotations*:Invasive ductal carcinoma/Invasive lobular carcinoma/Invasive micropapillary carcinoma/Invasive carcinoma with medullary features/ Invasive carcinoma, type cannot be determined |
| No distant metastasis | pathologic_stage; I/IA/IB/II/IIA/IIB/III/IIIA/IIIB/IIIC | Clinical TMN stage at II~III (early stage) | pathologic_stage: II/IIA/IIB/III/IIIA/IIIB/IIIC |
| Clinical TMN stage at I~III (early stage) | pathologic_stage: I/IA/IB/II/IIA/IIB/III/IIIA/IIIB/IIIC | | |
| High risk of recurrence (meet one of the following four conditions) | | | |
| 1) 4 or more positive axillary lymph nodes | ajcc_pathologic_n: N2/N2a/N3/N3a/N3b/N3c | | |
| 2) The tumor size is at least 5 cm | ajcc_pathologic_t: T3/T3a/T4/T4b/T4d | | |
| 3) Grade 3 defined as at least 8 points on the Bloom Richardson scoring system | N/A | | |
| 4) Ki-67≥20% | PAM50 and Claudin-low (CLOW) Molecular Subtype*:<br>LumB | | |

**c**

| NCT02675231<br>(the No.5 trial) | |
|---|---|
| **Eligible criteria of the clinical trial** | **Corresponding clinical information of TCGA patients** |
| 18 Years and older (Adult, Older Adult) | (TCGA-BRCA patients are all 18 years old and above) |
| Female | gender: Female |
| Postmenopausal status due to surgical/natural menopause or chemical ovarian suppression | menopause_status: Post |
| HR+ | er_status_by_ihc: Positive<br>OR<br>pr_status_by_ihc: Positive |
| HER2+ | her2_fish_status: Positive<br>copy number: Greater than or equal to 9<br>OR<br>her2_ihc_score: 3+<br>copy number: Greater than or equal to 9<br>OR<br>her2_status_by_ihc: Positive<br>copy number: Greater than or equal to 9<br>OR<br>her2_fish_status: [Not Evaluated]<br>her2_status_by_ihc: Positive/[Not Evaluated]/Equivocal/Indeterminate<br>copy number: Greater than or equal to 9 |
| Locally recurrent or metastatic disease | Metastatic:<br>ajcc_pathologic_m: M1<br><br>High risk of recurrence*:<br>ajcc_pathologic_n: N2/N2a/N3/N3a/N3b/N3c<br>OR<br>ajcc_pathologic_t: T3/ T3a/T4/T4b/T4d<br>OR<br>PAM50 and Claudin-low (CLOW) Molecular Subtype*: LumB |

*PAM50 and Claudin-low (CLOW) Molecular Subtype and Histology Annotations information are cited from the molecular analysis study of TCGA breast cancer histologic types[65]; Since TCGA clinical information doesn't include Ki67-related data, the LumB type was utilized in place of the condition "Ki-67 greater than or equal to 20%". This subtype represents individuals with a similar characteristic, i.e., Ki-67 greater than or equal to 15%.

*TCGA does not record the local recurrence status, so it is replaced by high risk of recurrence, presenting as the three listed conditions.

*TCGA staging (TMN staging) for the number of lymph nodes (N stage, ajcc_pathological_n), tumor size (T stage, ajcc_pathological_t), and cancer metastasis (M stage, ajcc_pathological_m) is based on the eighth edition of the AJCC manual.

*Other eligible criteria, such as the patient's systemic treatment history, ECOG physical status, etc., are not recorded in the TCGA clinical information or the number of records is too small to include.

**Supplementary Table 1 | The matched screening conditions for patients enrolled in simulated trials. a,** for No.1 clinical trial (NCT02246621) and No.2 clinical trial (NCT02079636). **b,** for No.3 clinical trial (NCT03155997) and No.4 clinical trial (NCT02513394). **c,** for No.5 clinical trial (NCT02675231).
