## Supplementary Table 2 for "A Genome-Informed Functional Modeling Approach to Evaluate the Responses of Breast Cancer Patients to CDK4/6 Inhibitors-Based Therapies and Simulate Real-World Clinical Trials"

### The baseline characters of No.3 and No.4 clinical trials

| | Simulated No.3 Clinical Trial (NCT03155997) n=197 | | Simulated No.4 Clinical Trial (NCT02513394) n=361 | |
|---|---|---|---|---|
| | n | % | n | % |
| **Gender** | | | | |
| Female | 195 | 98.98% | 358 | 99.17% |
| Male | 2 | 1.02% | 3 | 0.83% |
| **Age** | | | | |
| 20-30 | 1 | 0.51% | 4 | 1.11% |
| 31-40 | 20 | 10.15% | 25 | 6.93% |
| 41-50 | 38 | 19.29% | 78 | 21.61% |
| 51-60 | 58 | 29.44% | 96 | 26.59% |
| 61-70 | 49 | 24.87% | 94 | 26.04% |
| 71-80 | 22 | 11.17% | 45 | 12.47% |
| 81-90 | 9 | 4.57% | 19 | 5.26% |
| **Ethnicity** | | | | |
| American Indian or Alaska Native | 0 | 0.00% | 0 | 0.00% |
| Asian | 14 | 7.11% | 19 | 5.26% |
| Black or African American | 26 | 13.20% | 43 | 11.91% |
| White | 135 | 68.53% | 259 | 71.75% |
| Not reported | 22 | 11.17% | 40 | 11.08% |
| **AJCC pathologic stage** | | | | |
| I | 17 | 8.63% | 0 | 0.00% |
| II | 79 | 40.10% | 257 | 71.19% |
| III | 101 | 51.27% | 104 | 28.81% |
| IV | 0 | 0.00% | 0 | 0.00% |
| X | 0 | 0.00% | 0 | 0.00% |
| Not reported | 0 | 0.00% | 0 | 0.00% |
| **Pathologic type at primary diagnosis** | | | | |
| Infiltrating duct carcinoma, NOS | 143 | 72.59% | 252 | 69.81% |
| Lobular carcinoma, NOS | 42 | 21.32% | 89 | 24.65% |
| Infiltrating duct and lobular carcinoma | 4 | 2.03% | 7 | 1.94% |
| Infiltrating duct mixed with other types of carcinoma | 4 | 2.03% | 4 | 1.11% |
| Others | 4 | 2.03% | 6 | 1.66% |
| **Ki-67** | | | | |
| <15% | 112 | 56.85% | 237 | 65.65% |
| >=15% | 85 | 43.15% | 124 | 34.35% |

**Supplementary Table 2 | The baseline characteristics of the simulated No.3 and No.4 trials**
